# Quickest Way to Less Headache Days: an operational research model and its implementation for Chronic Migraine

**DOI:** 10.1101/2022.03.29.22273147

**Authors:** Irene Lo, Pengfei Zhang

**Affiliations:** Department of Neurology, Rutgers Robert Wood Johnson Medical School, New Brunswick, New Jersey, USA; Department of Management Science & Engineering, Stanford University, Stanford, California, USA

**Keywords:** Operations Research, Chronic Migraine, Headache, Medication Selection, Medical Decision Research, Markov Decision Process, Multi-armed bandit process

## Abstract

**Background:** Choosing migraine prevention medications often involves trial and error. Operations research methodologies, however, allow us to derive a mathematically optimum way to conduct such trial and error processes.

**Objective:** Given probability of success (defined as 50% reduction in headache days) and adverse events as a function of time, we seek to develop and solve an operations research model, applicable to any arbitrary patient, minimizing time until discovery of an effective migraine prevention medication. We then seek to apply our model to real life data for chronic migraine prevention.

**Methods:** We develop a model as follows: Given a set of D many preventive medications, for drug i in D, we describe the likelihood of reaching 50% headache day reduction over the course of time, (*t*_*i*,1_ ≤ *t*_*i*,2_ ≤ …) by probability (*p*_*i*,1_ ≤ *p*_*i*,2_ ≤ …). We additionally assume a probability of adverse event(*q*_*i*,1_ ≤ *q*_*i*,2_ ≤ …). We then solve for a sequence of prescription trials that minimizes the expected time until an effective drug is identified.

Once we identify the optimal sequence for our model, we estimate *p, t* and *q* for topiramate and OnabotulinumtoxinA based on the FORWARD study by Rothrock et al as well as erenumab data published by Barbanti et al. at IHC 2019.

**Results:** The solution for our model is to order the drugs by probability of efficacy per unit time. When the efficacy of each drug *i* is known only for one period *t*_*i*_ and there are no adverse effects, then the optimum sequence is to administer drug *i* for *t*_*i*_ periods in decreasing order *p*_*i*_/*t*_*i*_. In general, the optimum sequence is to administer drug *i* for *t*_*i,k*\*_ periods in decreasing order of the Gittins index *σ*_*i,k*\*_

Based on the above data, the optimum sequence of chronic migraine prevention medication is a trial of erenumab for 12 weeks, followed by a trial of OnabotulinumtoxinA for 32 weeks, followed by a trial of topiramate for 32 weeks.

**Conclusion:** We propose an optimal sequence for preventive medication trial for patients with chronic migraine. Since our model makes limited assumptions on the characteristics of disease, it can be readily applied also to episodic migraine, given the appropriate data as input.

Indeed, our model can be applied to other scenarios so long as probability of success/adverse event as a function of time can be estimated. As such, we believe our model may have implications beyond our sub-specialty.

**Trial Registration:** Not applicable.

## Introduction

Migraine has an approximate global prevalence of 14.7%. ^1^ Unfortunately, the process of finding an effective prevention therapy for patients is often largely trial and error process.^2, 3^ Medication trial failures frequently cause frustrations and a sense of helplessness among migraine sufferers.^3, 4^ Therefore an expedient way of determining effective prevention therapy can significantly benefit our patient population.

Operations research (OR) is the study of optimal decision-making using mathematical models. In healthcare, OR has been used to inform clinical trial designs and medical decision making in chronic diseases. ^5-7^ For example, OR models have been used to propose selections of chemotherapy combinations for clinical trials in oncology, to identify optimal timing and criteria for cancer screening, and to inform the optimal criteria for initiating medications such as statin in type 2 diabetics or highly active antiretroviral therapy in HIV patients. ^8-10^ OR models can also be used to support surgical decision-making, determining when to surgically correct abdominal aortic aneurysm or to receive kidney transplantation. ^11, 12^ To the best of our knowledge, however, operations research methods have not been applied to medication selection in headache.

The aim of this paper is to propose an operations research model for migraine medication selection. We assume that given any randomly selected patient one can estimate the probability of success for individual prevention medications as a function of time. We then solve our model for the optimal sequence of medication trial, minimizing time for the discovery of an effective treatment. As a proof of concept, we utilize published data to demonstrate that such a model can be implemented for real world decision-making purposes in chronic migraine prevention.

(Of note, our project was presented as a poster presentation during the American Headache Society’s June 2020 Virtual Meeting.)

## Methodology

Our methodology and results outline our operations research model for the clinician and layperson. We have included an in depth technical methodological portion for the technical reader in appendix A. Key proofs of our result are also included in appendix A.

### Setting up the model

For modeling purposes, we define successful/effective treatment response as 50% improvement in headache days. This value is not intrinsic to our model but is intrinsic to the input data. We assume that given any two medications, their respective probabilities of success are statistical independence. We assume that any intolerable adverse event would be discovered before observing 50% improvement in headache days. Finally, only one medication is allowed to be tried at a time.

Our model is as follows:

Given a set of D many preventive medications, for drug *i* in D, we describe the likelihood of reaching 50% headache day reduction over the course of time, (*t*_*i*,1_ ≤ *t*_*i*,2_ ≤ …) by probability (*p*_*i*,1_ ≤ *p*_*i*,2_ ≤ …). We additionally assume a probability of adverse event (*q*_*i*,1_ ≤ *q*_*i*,2_ ≤ …). We then solve for a sequence of prescription trials that minimizes the expected time until an effective drug is identified.

Topiramate, Botulinum Toxin (OnabotulinumtoxinA), and CGRP antagonists are the only medications widely accepted to have evidenced based efficacy in chronic migraine prevention.^13,14^ We use published 50% responder rates for week 12 and week 32 as well as adverse event rates from the FORWARD study as estimates for (*p*_*i*,1_≤ *p*_*i*,2_ ≤ …). And (*q*_*i*,1_≤ *q*_*i*,2_ ≤ …). for topiramate and OnabotulinumtoxinA. ^15^ We use 50% responder rates from Barbanti and colleagues’ retrospective study to estimate probability of real world efficacy as a function of time for erenumab. ^16^ We have decided against using clinical trial data due to methodological concerns; galcanezumab and fremanezumab are therefore not included in this study. (See Limitation Section.) We estimate adverse event for erenumab based on package insert. ^17^ Our input data for the model is summarized in Table 1.

(A quick note on notation: The set D in our case represents the set of three medications: OnabotulinumtoxinA, topiramate, erenumab. The subscript *i* therefore represents one of these medications. The second subscript *k*, represents the epoch of time under consideration. The notation P(x) represents probability of x and E(x) represents expected value of x, in accordance with traditional mathematics parlour. The asteroid (*) next to a variable is used to denote optimum solution.)

### Theoretical Results

Our model a case of the multi-armed bandit problem in operations research. Solution sto this class of problem can be solved through calculation and identification of a ratio called the Gittins Index at every decision point.^23^ In other words, index ratios are calculated for all possible choices at every decision point; one then identifies the next optimum decision by picking the decision with the highest index. We denote the calculation of these indices as σ_*i,k*_. We denote the Gittins index as σ_*i,k*\*_

The solution for our model is to order the drugs by probability of efficacy per unit time. Let us first consider an unrealistic but simplified scenario to our problem: When the efficacy of each drug *i* is known only for one period *t*_*i*_, then the optimum sequence is to administer drug *i* for *t*_*i*_ periods in decreasing order of *p*_*i*_/*t*_*i*._ For example, under this simplified scenario, suppose a hypothetical antiepileptic has a 90% probability of being effective (let’s call this *p*_*AED*_) only by the end of 3 months (*t*_*AED*_) whereas a hypothetical CGRP mab has a probability of 50% (p_CGRP_) only by the end of 1 month (*t*_CGRP_). Then it is wise to try the hypothetical CGRP first for 1 month followed by trying the antiepileptic for 3 months if the goal is to discover an effective medication as quickly as possible. Note that in this simplified scenario we assume that one does not see any intermediate benefit between month 0 and month 3 for the antiepileptic nor any intermediate benefit between day 0 and day 1 for the CGRP. This, of course, is not realistic. (The proof of this result is under the Special Case section in Appendix A.)

Our problem is of course more complicated than the simplified version above and a generalized, and therefore more realistic, solution to the above can be derived: the optimum sequence is to administer drug *i* for *t*_*i,k\**_ periods in decreasing order of the index σ_*i,k*\*_, where

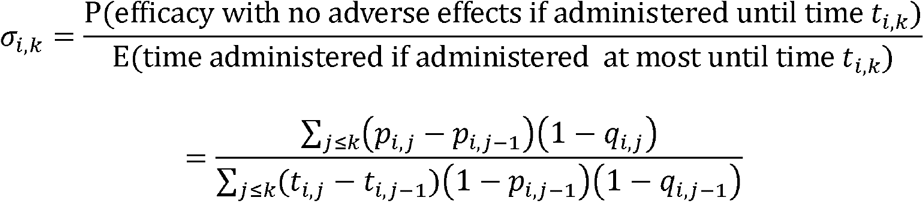

is the probability of efficacy per unit time if the drug were administered until time *t*_*i,k*_, and

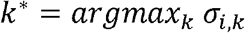

is the number of time periods that maximizes the efficacy per unit time of the drug. Note that in order to determine which drug to try next, the indices for the trialed drug *i* need to be recalculated after trying it for time *t*_*i,k\**_. (Proof of the above results are included in Appendix A.)

In other words, σ_*i,k*_ represents a generalized version of *p*_*i*_/*t*_*i*._ The numerator of σ_*i,k*_ represents the probability of the drug efficacy if administered until time *t*_*i,k*._ The denominator σ_*i,k*_ of represents the time the drug is administered to the patient before time *t*_*i,k*_ taking into account that this time may be shorter than *t*_*i,k*_ if the drug is successful earlier in the process.

To calculate the index for individual drugs, we must first find the optimal medical trial time frame *t*_*i,k\**_, represented in *k*, by picking *k\** such that it maximizes σ_*i,k*_. This is the calculation of the Gitten’s index. Once these selections are obtained, then we simply rank them in decreasing orders of σ_*i,k*\*_, trying each drug for *t*_*i,k\**_ periods.

### Application of model to chronic migraine data

Based on FORWARD as well as Barbanti study data, we calculated the above indices for the first iteration and present the results in Table 2. (Here, σ_*i*k_ represents the index for each medication at specific time k for drug i.) Input probabilities are adjusted to ensure probabilities are increasing over time, this is done by simply taking the maximum probability of efficacy up to each specific time frame.

Note that regardless of potential erenumab trial duration, the resulting index is larger than the largest indices for both of the other drugs. Therefore it is optimal to try erenumab first for the full 12 weeks. As such, one optimal sequence of chronic migraine prevention medication is:

1. A trial of erenumab for 12 weeks
2. If the above fails, then proceed with a trial of OnabotulinumtoxinA for 32 weeks
3. If the above fails, then proceed with a trial topiramate for 32 weeks.

Since estimates for OnabotulinumtoxinA and topiramate are based on a lower and upper bound based on our input data (the FORWARD study contains these values), it is possible that the following alternative sequence is also optimal:

1. A trial of erenumab for 12 weeks
2. If the above fails, then proceed with a trial of OnabotulinumtoxinA for 12 weeks
3. If the above fails, then proceed with a trial of topiramate for 12 weeks
4. If the above fails, switch back to a trial of OnabotulinumtoxinA for 20 further weeks
5. If the above fails, switch back to a trial of topiramate for 20 further weeks.

This alternative optimal solution is, of course, only a theoretical one given that switching between OnabotulinumtoxinA and topiramate would be logistically difficult and clinically peculiar.

### Instrumentation

Our model is not a simulation and therefore the results can be calculated by hand. We, have, however, conducted our calculations through the use of Microsoft Excel.

## Discussion

In this proof of concept study, we frame the process of migraine preventive medication selection as an operations research problem. We then outline a methodology for the construction of an operations research model, its mathematical solution, and its real life implementation. We believe such a perspective will be invaluable to clinical practice. Applications of operations research to medical decision making/clinical trial design have relied mostly on Markov Decision Process (MDP). ^18, 19^ Traditionally, medical decision-making MDP models are personalized, having the benefit of producing optimizing policy through individual-specific information as well as treatment responses. ^9-12^ However, solutions to personalized MDP models can be computationally demanding, requiring large amount of data as input and are often difficult to interpret for practitioners. These limitations have prevented widespread use of personalized MDP models clinically. ^10, 20^ As an alternative to personalized MDP, our paper utilizes a multi-armed bandit process for medication selection. A multi-armed bandit process is a special case of MDP well studied in OR and utilizes an “index” method. Our method is based on an indexing method proposed previously for evaluation of sequential clinical trial design. ^5, 6^ We are unable to find a prior instance of utilization of this method in clinical decision-making for medication selection.

We want to be very clear, however, that our model is not intended to be utilized to the exclusion of all other patient centered concerns such as comorbidities, access, or drug-drug interactions. Indeed, mathematical modeling is not a substitute for good doctoring. For example, obviously we do not recommend practitioners initiate topiramate if the patient has a history of nephrolithiasis, nor do we think that one should start off with erenumab as a treatment should the patient has a history of severe hypertension or severe constipation. Furthermore, mathematical modeling is not a panacea for addressing medication overuse headaches, obtaining insurance approved to prevention medication, avoiding medication interactions, assessing patient preference, and all other clinical decision making processes. Addressing these real-life concerns are beyond the scope of this paper. In other words, mathematical model, just like animal models in biological sciences, is meant to inform clinicians/scientists.

By using a multi-arm bandit process, our model allows for an important intuitive result: that we can estimate optimal sequence for prevention medication selection with the index *p*_*i*_/*t*_*i*_. This provides a clinically intuitive way of understanding medication optimization – prioritizing and select medications that maximizes probability of efficacy per unit of time. In accordance with our model, this index should be re-evaluated in real time and on each follow up visits to offer continual guidance for the next medication choice.

Although the focus of this paper is chronic migraine, our model and its mathematical solution can be readily translated to episodic migraine as long as probabilities for success/adverse events as a function of time can be estimated. The insight for optimizing the index *p*_*i*_/*t*_*i*_ can be directly translate to episodic migraine selection as well.

Finally, we believe that operations research models for medication selection should not be limited to our subspecialty. For example, one can imagine applying a similar approach to managing epilepsy and multiple sclerosis. Given that we make no assumption on the underlying disease process, our model and its solution may be directly applicable to a wide range of clinical medicine scenarios outside our discipline.

### Strength and Limitations

Although we defined success in our model as 50% improvement, this number is not intrinsic to our model. In other words, the mathematics of our model would work equally well for any other parameter so long as the input data contains efficacy data as a function of time for the same parameters. That is, the logic for our model can be carried over to efficacies defined as, for example, 25% improvement, or 75% improvement, or even 50% decrease in headache severity with the caveat that we have data on how these parameters change as a function of time given specific medications. Indeed our model is limited by the availability of real life data.

Our model’s assumptions are also its limitations: First, we assume mono-therapy in medication selection. While we believe this to be a reasonable assumption, we acknowledge that this is not often true in the real world. For example, it is not uncommon for patient to be prescribed topiramate while waiting for OnabotulinumtoxinA, resulting in eventual continuation of both OnabotulinumtoxinA and topiramate for prevention. Challenging our paradigm of monotherapy is also the open question whether synergism exists between OnabotulinumtoxinA and CGRP antagonists. Of course, one can argue that the quickest way for treating chronic migraine would be to simply provide OnabotulinumtoxinA, CGRP, and topiramate concurrently. This solution is possible but often not feasible in real life due to economic limitations and clinical considerations.

Our model also does not take into account how patient-specific factors affects efficacies of individual medications. This is, in part, due to the lack of data for patient-specific factors for various medications as a function of time. For example, an extension of our model may be able to account for efficacies of topiramate as a function of time when body mass index (BMI) is taken into consideration. However, we would then require multivariable data on topiramate efficacy as a function of both time *and* BMI.

Our most significant limitation is therefore the scarcity of available data for our model. Firstly, as a proof of concept paper we are not wedded to the idea of using only FORWARD nor Barbanti data for our model; the primary purpose of this paper is a methodological one after all. Secondly, we have decided to only include real-life data based on the critique that clinical trial responder rates may not translate directly to real word settings. This methodological concern is especially regrettable in regards to CGRP data: While Forderreuther et al as well as Tepper et al estimated 50% response rates of erenumab and galcanezumab as a function of time, these study were extrapolated from clinical trial, therefore limiting their application to this paper. ^21, 22^ Finally, although multiple real-life data exist for OnabotulinumtoxinA efficacy, apart from the FORWARD study we are unable to locate a study where 50% responder rate as a function of time is documented.

### Future directions

Our model’s limitations are sources for future directions: In order to apply our solution to real life decision-making, we require real life data in the form of responder rate as a function of time. Our model shows that these real-world data can significantly affect clinical decision-making. We hope that this paper will alert future researchers to the importance of real-world data, allowing for improvement in implementation of our method.

## Conclusions

An operation research model can be constructed for optimal medication decision-making in headache prophylaxis. We outline the methodology for such an operations research model and its empirical implementation. Although our model supports the sequence of erenumab, followed by OnabotulinumtoxinA, followed by a trial of topiramate, more data is needed to support this sequence as being optimal. Our model can be applied to episodic migraine should more real life data be made available. We believe a similar approach can be applied to other specialties.

### Article Highlights

- We outlined a methodology for the construction of an operations research model, its mathematical solution, and its real life implementation for selection of migraine prevention medications minimizing time until efficacy.
- One can estimate optimal sequence for prevention medication selection with the index *p*_*i*_/*t*_*i*_. (A generalized and more realistic version of the above index is the Gittins Index.) This provides a clinically intuitive way of understanding medication optimization – prioritizing and select medications that maximizes probability of efficacy per unit of time.

## Data Availability

All data produced in the present study are available upon reasonable request to the authors

## Abbreviations

(IHC): International Headache Congress
(OR): Operations Research
(HIV): Human Immunodeficiency Virus
(MDP): Markov Decision Process
(CGRP): calcitonin gene-related peptid

## Acknowledgements

Laura Hester Ph.D for feedback on our project.

## Appendix A

Proof of the key results

Let

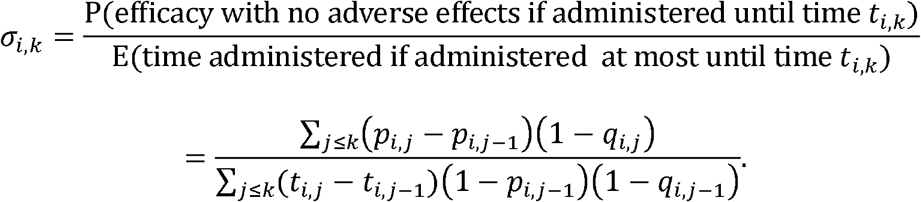

We show that the expected time to effective treatment is minimized using the following procedure:

1. Calculate σ_*i,k*_, for all arms *i* and periods *t*_*i,k*_.
2. Find the largest σ_*i,k*_ call this σ_*i,k\**_
3. Administer drug *i* for *t*_*i,k\**_ periods
4. If drug *i* is not effective after *t*_*i,k\**_ periods, recalculate σ_*i,k*_ for all subsequent time periods for arm *i* and repeat steps 2-4.

### Special Case

To illustrate why such a procedure should work, we first show that it minimizes the time to effective treatment when the efficacy of each drug *i* is known only for one period *t*_*i*_ and there are no adverse effects each drug. In this case

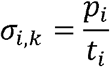

and we show that the optimal prescription sequence is to prescribe drugs in decreasing order of *p*_*i*_/*t*_*i*._

Suppose not, so there is some drug prescription sequence S that does better. Let us assume that the drugs are labeled so that the sequence S is in the order 1,2,…, *n*. Since the drugs are not in decreasing order of *p*_*i*_/*t*_*i*_, there are two drugs *i,j* such that *p*_*i*_/*t*_*i*_ > *p*_*j*_/*t*_*j*_ and drug *i* is prescribed after drug *j*. (In fact we can assume that drug *i* is prescribed right after drug *j*, so *j =i* − 1). Then

E [time to effective treatment with sequence S]

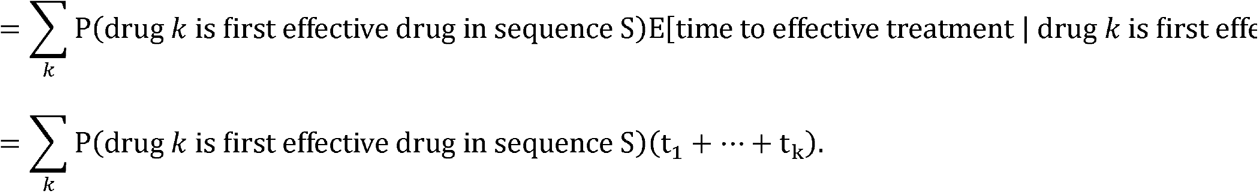

Let S’ be the sequence where the order of drugs *i* and *j* are swapped. We show that the expected time to effective treatment with S’ is strictly less than with S:

E [time to effective treatment with sequence S′]

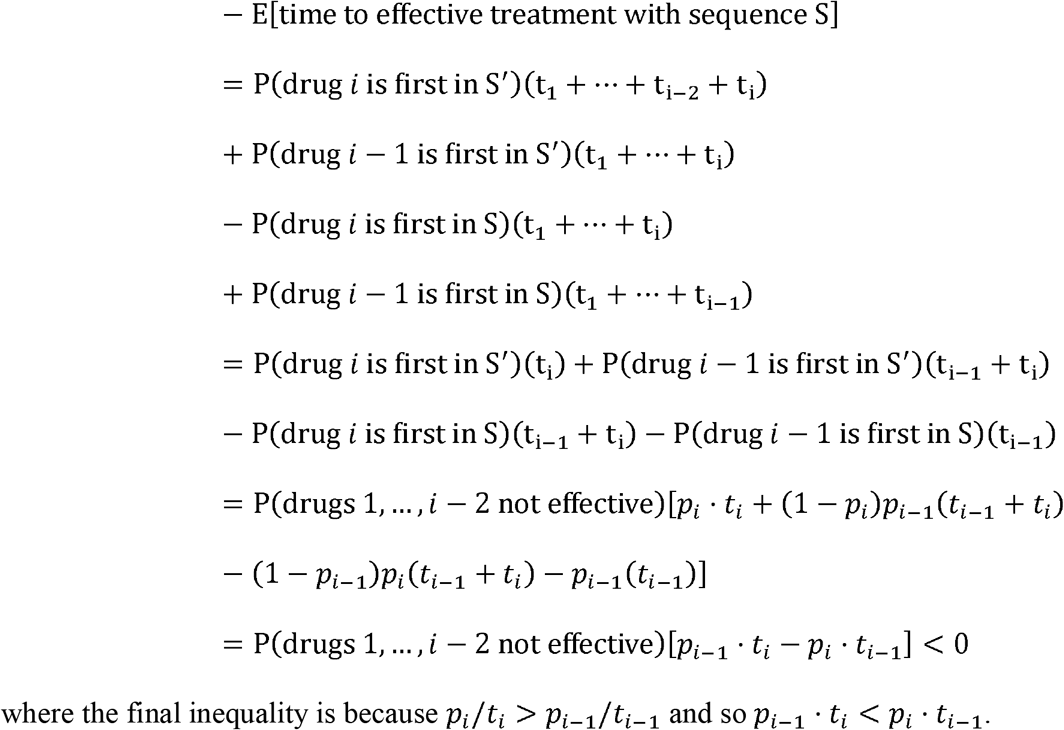

### General Case

We now show that when drug efficacy and probability of adverse effects are known for multiple periods the procedure outlined above minimizes expected time to effective treatment.

For each drug, we can think of taking one of two actions: either trying the drug for another week, or not trying the drug for another week. For every week we try the drug we pay a cost of 1. If we try the drug for another week and it is effective with no side effects, we receive a reward of R, otherwise we receive a reward of 0. Once the drug is effective or the patient exhibits adverse effects we receive no further rewards for trying the drug. The decision-maker must sequentially choose every week at most one drug to prescribe for that week, with the goal of minimizing the expected time until efficacy. For sufficiently large R this goal is the same as maximizing the total reward minus cost.

This kind of problem is precisely a **multi-armed bandit process**, a special case of a Markov decision process that is well-understood by the operations research literature. A general multi-armed bandit process is defined as follows. There are *n* arms, each arm *i* starting in a given state *x*_*i*_ For each arm *i*, we can take one of two actions: either pulling the arm this time period, or not pulling the arm this time period. If we pull the arm we receive some random reward *r*(*x*_*i*_) that is a function of the state of the arm, and the arm randomly transitions to a new state that is a function of the current state of the arm. The decision-maker must sequentially choose in every period which arm to pull, with the goal of maximizing the total reward. There is a well-known solution to the multi-armed bandit process, which was first formulated by J. C. Gittins and D. M. Jones and is given by the following:

### Theorem

The expected discounted reward obtained from a multi-armed bandit problem is maximized by pulling the arm with the greatest Gittins index:

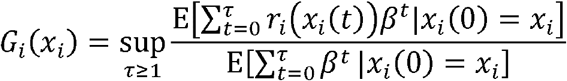

where τ is a past-measurable stopping time. ^23^

Let us rewrite the drug prescription problem as a multi-armed bandit problem. Let *T*_*i*_ = max_*k*_ *t*_*i,k*_ be the longest amount of time we can prescribe drug *i*. Each arm *i* (representing drug *i*) can be in one of the states {1, …, *T*_*i,*_*s*, −1}, where 1 represents the initial state, *s* represents the state when the drug is first successful, −1 represents the state where no more rewards are received, and *k* ∈ {1,2, …, *m*_*i*_} represents the number of times drug *i* has been tried. Arm *i* starts in the state ‘1’. From each state *k* ∈ {1, …, *T*_*i*_}, if arm *i* is pulled the ‘reward’ is −1 (i.e. paying a cost of 1 for each week the drug is tried). From each state *k* ∈ {*t*_*i*,1_,*t*_*i*,2_, …} and the state changes randomly according to the following probabilities: with probability *p*_*i,k*_ (1−*q*_*i,k*_)the drug is successful with no adverse effects and the state changes to *s*, with probability *q*_*i,k*_, there are adverse effects and the state changes to −1 and with probability (1 − *p*_*i,k*_) (1 − *q*_*i,k*_) the drug is not successful with no adverse effects and the state changes to *k* + 1 (or to state −1 if *k* = *m*_*i*_). From state *s* if arm *i* is pulled the reward is *R* and the state changes to −1, and from state −1 if arm *i* is pulled the reward is 0 and the state stays at −1. Finally, from all other states *k* ∈ {1, …, *T*_*i*_}\{*t*_*i*,1_,*t*_*i*,2_, …} the reward is −1 and the state changes to *k* +1. The goal is to maximize the expected total reward, which is given by

*R*(probability success) − (time until find successful drug with no adverse effects).

For sufficiently large *R* this is the same as minimizing the expected time until success. Our rewritten drug prescription problem is a special case of a multi-armed bandit problem where the reward is deterministically 0 after some finite time, and so the expected total reward is maximized by pulling the arm with the greatest Gittins index:

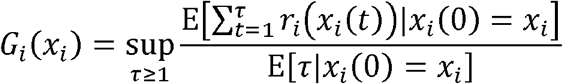

where *τ* is a past-measurable stopping time. In this case the only values of *τ* where we can receive positive reward (and hence the only values of *τ* we have to care about) are {*τ*_*i*,1_, *τ*_*i*,2_, …} where is the random stopping time *τ*_*i,k*_ corresponding to pulling the arm until the first time before *t*_*i,k*_ there is a success, and then pulling once more. Hence

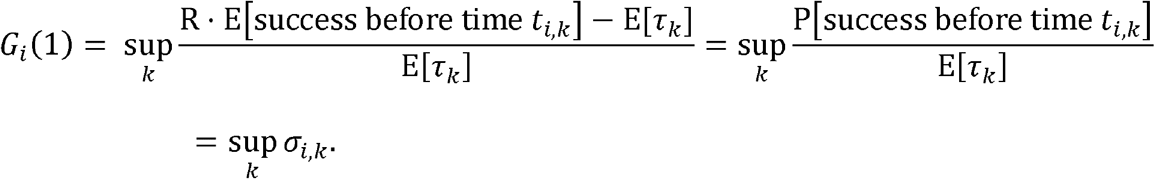

